# Vitamin D deficiency and toxicity across 2018 to 2022 in several cities of Ecuador: interrupted time series and a cross-sectional study

**DOI:** 10.1101/2023.09.08.23295127

**Authors:** Camilo Zurita-Salinas, Betzabé Tello, Iván Dueñas-Espín, Jeannete Zurita, William Acosta, Cristina Aguilera León, Andrés Andrade-Muñoz, José Pareja-Maldonado

## Abstract

**Objectives:** to identify differences in mean vitamin D concentrations in samples obtained from a private laboratory in the city of Quito, and to explore their relationship with the pre-pandemic and pandemic periods spanning from 2018 to 2022.

**Design:** A combination of an interrupted time series design and a retrospective cross-sectional approach

**Setting and participants:** The study involved 9,285 participants who had their 25-hydroxyvitamin D (25(OH)D) levels tested at a well-known private laboratory in Quito, Ecuador, from 2018 to 2022.

Primary and secondary outcome measures: The 25(OH)D levels were analyzed and assessed for correlations with both age and the year in which the measurements were taken.

**Results:** The mean 25-hydroxyvitamin D (25(OH)D) level was 27.53 ng/mL (± 14.11). Approximately 64.58% of participants had insufficient levels, below 20 ng/mL, and 0.62% showed potential harm from excess 25(OH)D, with levels over 100 ng/mL. The analysis indicated a significant monthly increase of 0.133 units in 25(OH)D levels (p=0.006). However, the period after March 2020, compared to before, saw a non-significant decrease of 1.605 units in mean 25(OH)D levels (p=0.477).

**Conclusions:** The study’s findings indicate a significant prevalence of 25-hydroxyvitamin D (25(OH)D) deficiency, underscoring the necessity for preventative measures. Nevertheless, the rise in cases of vitamin D toxicity is concerning, emphasizing the importance of prudent vitamin D supplement prescriptions and public education against self-medication. For efficient resource allocation and targeting those with higher risks, it may be advantageous to concentrate vitamin D testing on specific population groups.

## Introduction

Vitamin D, a prohormone, has a crucial influence on a multitude of biological processes, encompassing immunological responses, phosphocalcic metabolism, and detoxification. Recent revelations regarding the diverse functions of vitamin D have engendered escalating interest among the scientific community, fostering an ongoing commitment to further research in this realm. The intricate regulation of vitamin D is governed by a complex interplay of genetic and environmental factors, underscoring the multifaceted nature of its mechanisms [1].

The primary source of vitamin D in humans stems from the interaction of UVB radiation with the skin during sunlight exposure. When the skin is exposed to ultraviolet radiation, a photochemical reaction occurs; cleaving carbon bonds in the precursor of vitamin D, 7-dehydrocholesterol, to form pre-vitamin D2. Subsequent temperature-dependent molecular rearrangements facilitate the production of active vitamin D, which can be stored within the body for several months due to its liposoluble nature[2]. In addition to sunlight, vitamin D can also be obtained from external sources such as certain foods and supplements[1].

Defining the optimal level of 25(OH)D, the major circulating form of vitamin D, remains a topic of ongoing debate[3]. Some studies have highlighted a potential overestimation of vitamin D deficiency prevalence in the population, fueling controversy over the necessity of supplementation in healthy individuals[3]. Notably, The Endocrine Society defines deficiency as <20 ng/ml, insufficiency as 21-29 ng/ml, and optimal levels as >30 ng/ml, while the Institute of Medicine (Health and Medicine Division of the National Academies) considers deficiency as <12 ng/ml, insufficiency as 12-20 ng/ml, and optimal levels as >20 ng/ml. Furthermore, The American Association of Clinical Endocrinologists defines deficiency as < 30 ng/ml and optimal levels as 30-50 ng/ml[4]. Globally, an estimated 15.7% of the population suffers from a vitamin D deficit, with South America exhibiting a higher prevalence of 34.75%[4,5]. No previous studies in Ecuador have considered altitude and the pandemic. Quito, Ecuador’s capital, sits at 2850 meters above sea level.

During the COVID-19 pandemic, several studies have documented significant lifestyle changes among individuals, including modifications in dietary patterns, reduced participation in outdoor activities, and a decline in physical exercise[6]. These lifestyle shifts have the potential to impact vitamin D synthesis, as they may serve as contributing factors[6,7].

Vitamin D deficiency has been linked to acute and chronic diseases that affect not only the skeletal system but also other physiological systems. Timely treatment for individuals with vitamin D deficiency can potentially improve their quality of life and reduce associated health risks[8].

The objective of this study was to identify differences in mean vitamin D concentrations in samples obtained from a private laboratory in the city of Quito, and to explore their relationship with the pre-pandemic and pandemic periods spanning from 2018 to 2022.

## Methods

### Design

A combination of an interrupted time series design and a retrospective cross-sectional approach was employed in this study to assess shifts in population-level 25-hydroxyvitamin D (25(OH)D) status and associated toxicity in Ecuador, pre - and post-onset of the COVID-19 pandemic. Data were collated at multiple time points through cross-sectional measures of previously collected data prior to and following the pandemic’s onset. This design facilitated the appraisal of long-term trends and the potential influence of the pandemic on 25-hydroxyvitamin D (25(OH)D) levels and associated toxicity.

### Settings

This study was conducted primarily in a large private laboratory in Quito, Ecuador. A diverse cohort of patients was included, most of whom had their 25-hydroxyvitamin D (25(OH)D) levels tested at the request of their physicians. These people came from various cities, primarily from Quito, Ibarra, Ambato, and Santo Domingo. Data from 2018 to 2022 were analyzed, with the exception that for the year 2022 only data from January were included.

### Study population and sample size

The study population comprised people who had their 25-hydroxyvitamin D (25(OH)D) levels tested at a large private laboratory in Quito, Ecuador, between the years 2018 and 2022. A vast majority of these tests were performed at the request of the patients’ physicians. Patients from various cities such as Quito, Ambato, Ibarra, and Santo Domingo contributed to the diversity of the sample. The total sample size was 9,285. By the year 2022, the sample included only those people whose vitamin D levels were tested in January. Additionally, we performed a secondary analysis in a subsample of 919 patients in which we collected information about chronic diseases and medications in order to calculate the prevalence of inadequate levels of vitamin D in people with and without chronic diseases.

### Data collection procedures

There is a consensus among experts regarding the use of serum/plasma 25(OH)D concentration as the preferred method to evaluate vitamin D status. It is widely agreed that this measurement reflects the combined contributions of diet and dermal synthesis. The choice of 25(OH)D is justified by its advantageous characteristics, including its extended half-life of 15 days, relative stability, abundant presence in the blood, and responsiveness to recent endogenous vitamin D production and exogenous intake from diet or supplements[1,9]. The determination of 25-hydroxyvitamin D Total in human plasma was assessed using the ELFA (Enzyme Linked Fluorescent Assay) technique. The measurements were performed using the VIDAS laboratory machine, employing reagents manufactured by Biomerieux. Following the completion of the assay, the results were automatically analyzed by the computer.

The instrument automatically calculated the results using stored calibration curves based on a 4-parameter logistics model. The results were expressed in either ng/mL or nmol/L.

The VIDAS 25 OH Vitamin D TOTAL measurement range spanned from 8.1 ng/mL to 126.0 ng/mL. Results below the lower limit of the measurement range were reported as “< 8.1 ng/mL,” while values above the upper limit were reported as “> 126.0 ng/mL.”. In this study, the reference value of < 20ng/mL was adopted to classify laboratory samples as “inadequate levels”, encompassing values indicating deficiency and insufficiency. Furthermore, values exceeding 100 ng/mL were considered “suggestive toxicity”.

### Variables

The main variables of interest in this study were serum concentrations of 25-hydroxyvitamin D [25(OH)D], age, and the year of measurement. Vitamin D levels were classified as ’deficiency’, ’insufficient’ and ’sufficient’ based on recognized clinical thresholds. We further classified “Suggestive toxicity” when the values were >100 ng/mL, and “Inadequate levels” when the values were deficient or insufficient. Age was stratified into four categories: ‘Children and adolescents’, ‘Young adults’, ‘Middle-aged adults’ and ‘Older adults’. The year of measurement was used to compare vitamin D levels before and after the start of the COVID-19 pandemic. We also considered two outcome variables: ‘Suggestive toxicity’ and ‘Inadequate levels’.

### Statistical analysis

Descriptive statistics were first calculated to summarize the basic characteristics of the data set, including means and standard deviations for continuous variables, and frequencies and percentages for categorical variables. We stratified the analyzes by age group and by year of measurement to assess temporal trends in 25-hydroxyvitamin D (25(OH)D) levels. Chi-square tests were used to examine differences in the distribution of categorical variables between different groups. For continuous variables, independent samples t tests or Mann-Whitney U tests were used, as appropriate. A p-value of less than 0.05 was considered statistically significant. To further explore the relationships between age, year of measurement, and 25-hydroxyvitamin D (25(OH)D) levels, cross-tabulations were performed. These analyzes were used to identify the prevalence of 25-hydroxyvitamin D (25(OH)D) deficiency, insufficiency, sufficiency, suggestive toxicity, and inadequate levels within each age group and year. Results are reported as mean ± standard deviation (SD) for continuous variables and number (percent) for categorical variables, unless otherwise stated.

Two secondary analyses were performed: First, within a subsample of participants to investigate possible selection bias in the study results. The association between self-reported illnesses and inadequate 25-hydroxyvitamin D (25(OH)D) levels was assessed using a chi-square test. Medical conditions were self-reported using the personal data form, and this information was integrated into the laboratory’s database, along with data on medication use and comorbidities. Second, a secondary analysis was performed to compare 25-hydroxyvitamin D (25(OH)D) levels between cities located at different altitudes: Quito, Ambato, and Ibarra, located above 2,500 masl, and Santo Domingo, located below 625 masl. The chi-square test was used to examine the association between city location and inadequate vitamin D levels.

To examine the potential change in average 25-hydroxyvitamin D (25(OH)D) concentration before and after March 2020 (pre-pandemic and pandemic periods), a multiple linear regression analysis was employed. The independent variables in the analysis included the passage of time (measured in months), a binary variable indicating the period after March 2020, and an interaction term combining these two variables. The dependent variable was the average monthly 25-hydroxyvitamin D (25(OH)D) concentration. The total sample size for the regression analysis was 49.

For the statistical analysis, a Poisson regression model was utilized to determine the prevalence ratios for two health outcomes, namely toxicity and inadequate levels of 25-hydroxyvitamin D (25(OH)D), over a four-year period (2018-2021). For each outcome, the year of observation was included as a categorical independent variable, and prevalence ratios were estimated in relation to the base year of 2018. Observations from the year 2022 were excluded from the analysis.

All statistical analyzes were performed using Stata 15.1 (Stata Corp. 2015. Stata Statistical Software: Release 14. College Station, TX: StataCorp LP). A significance level of p<0.05 was considered statistically significant, indicating the presence of significant differences between the groups.

### Ethical issues

The study protocol (CEISH 659-2022) received ethical approval from the Ethics Committee for Research in Human Beings of the Pontifical Catholic University of Ecuador, following an expedited review process.

## Results

Descriptive results from the total sample are shown in **Table 1**. In this study population of n=9286, most participants were female (74.65%). The mean age of the population was 51.58 years (± 23.00), and the age range varied from 1-2 years to 100-130 years. The mean concentration of 25-hydroxyvitamin D (25(OH)D) was 27.53 ng/mL (± 14.11). Regarding 25-hydroxyvitamin D (25(OH)D) levels, 64.58% of the participants had inadequate levels, defined as a concentration of 25-hydroxyvitamin D (25(OH)D) below 20 ng/mL. In addition, suggestive toxicity, indicating potential harm due to excess 25-hydroxyvitamin D (25(OH)D), was present in 0.62% of the population, with a 25(OH)D concentration limit greater than 100 ng/mL. There was missing information because some participants did not provide their sex. The presented means and standard deviations provide more information about the age distribution and 25-hydroxyvitamin D (25(OH)D) concentration within the study population.

**Table 1:** Demographics, 25-hydroxyvitamin D (25(OH)D) Levels, and Inadequate and Suggestive Toxicity of 25-hydroxyvitamin D (25(OH)D) of the Study Population (N=9286)

| Variable | Statistics |
| --- | --- |
| **Sex** |  |
| Female, n (%) | 6932 (74.65%) |
| Male, n (%) | 2318 (24.96%) |
| Not registered, n (%) | 36 (0.39%) |
| **Mean Age (± SD)** | 51.58 years (± 23.00) |
| **Age Range (Years)** |  |
| 1-2, n (%) | 34 (0.37%) |
| 3-5, n (%) | 93 (1.01%) |
| 6-11, n (%) | 275 (2.98%) |
| 12-17, n (%) | 440 (4.76%) |
| 18-65, n (%) | 5630 (60.92%) |
| 66-79, n (%) | 1685 (18.23%) |
| 80-99, n (%) | 1070 (11.58%) |
| 100-130, n (%) | 14 (0.15%) |
| **Mean 25-hydroxyvitamin D (25(OH)D)**<br>**Concentration (± SD)** | 27.53 ng/mL (± 14.11) |
| **Inadequate 25-hydroxyvitamin D (25(OH)D)**<br>**Level** |  |
| Yes, n (%) | 5997 (64.58%) |
| **Suggestive Toxicity** |  |
| Yes, n (%) | 58 (0.62%) |
| *Notes:* |  |
| 1. *Inadequate 25-hydroxyvitamin D (25(OH)D) Level* was defined as having a concentration of 25-hydroxyvitamin D (25(OH)D) less than 20 ng/mL. |  |
| 2. *Suggestive Toxicity* was defined based on the presence of symptoms indicating potential harm or risk due to excess 25-hydroxyvitamin D (25(OH)D), as ascertained by medical evaluation, and when the concentration of 25-hydroxyvitamin D (25(OH)D) exceeded 100 ng/mL. |  |
| 3. Participants who did not disclose their gender are listed as 'Not Disclosed' under the 'Sex' category. |  |
| 4. Age is categorized into ranges for easier data interpretation. |  |
| 5. The means for age and 25-hydroxyvitamin D (25(OH)D) concentration are presented with their respective standard deviations. |  |

Additionally, regarding the two secondary analyses, First, in the subsample analysis, no significant differences were found in inadequate 25-hydroxyvitamin D (25(OH)D) levels between those without self-reported diseases (63%), those without any medication (64%), those with any self-reported disease (64%), and those taking any medication (61%) (**S1 Table**). Inadequate levels were considered based on specific criteria related to Vitamin D status. These findings suggest that the results of our study are more generalizable to the general population from which the sample was drawn, since there does not seem to be a selection bias related to the presence of diseases. Furthermore, when comparing 25-hydroxyvitamin D (25(OH)D) levels between cities located at different altitudes: Among the participants from Quito, Ambato and Ibarra, 64.18% had inadequate levels, while 52.74% of the participants from Santo Domingo had inadequate levels (chi-square = 18.1854, p < 0.001). These findings suggest that altitude may play a role in 25-hydroxyvitamin D (25(OH)D) status, as people residing at higher altitudes are more likely to have inadequate levels.

Regarding the evolution of concentration of 25-hydroxyvitamin D (25(OH)D) across years, (**S2Table**) provides an overview of 25-hydroxyvitamin D (25(OH)D) levels, suggestive toxicity, and inadequate levels in different years and age groups. In 2018, a significant difference in the prevalence of deficiency was observed between age groups, with the highest proportion in children and adolescents (42.9%), followed by young adults (43.0%), middle-aged adults (20.4%) and older. adults (33.0%). Similarly, in 2021 a significant difference was found, with the highest proportion of deficiency in children and adolescents (29.1%), followed by older adults (26.1%), young adults (28.0%) and middle-aged adults (21.6%). Regarding inadequate levels, the prevalence varied between years and age groups, with the highest figures being observed in 2021 (2,695) and in the group of middle-aged adults (1,099). Suggestive toxicity was identified in a small number of cases, mainly in older adults. In general, there was a significant association between the year and the prevalence of deficiency and inadequate levels of 25-hydroxyvitamin D (25(OH)D), indicating variations over time.

We can observe the trend of monthly average plasma of 25-hydroxyvitamin D (25OHD) concentration during the pre-pandemic and pandemic periods. During the pandemic period, although the values of 25-hydroxyvitamin D (25OHD) remain inadequate, there is an evident increase in the monthly average concentration of 25-hydroxyvitamin D (25OHD). It increases from 2.4 ng/mL in 2018 to 2022 (Figure 1).

**Figure 1.**
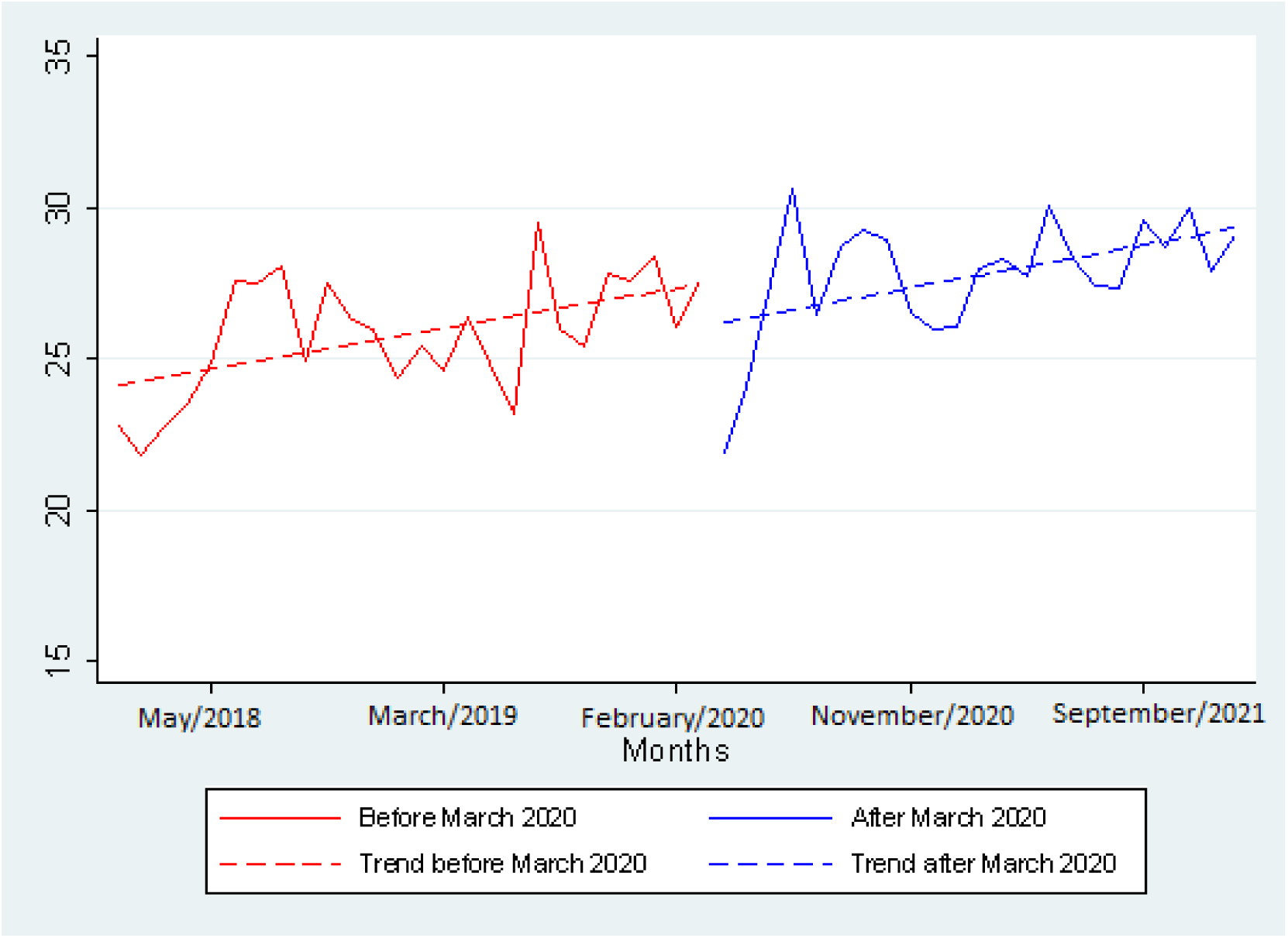
Monthly mean 25-hydroxyvitamin D (25OHD) concentration pre-pandemic and pandemic period.

The results of the regression analysis (**Table 2**) revealed that there was a statistically significant average monthly increase in 25-hydroxyvitamin D (25(OH)D) concentration of 0.133 units (p=0.006). The period after March 2020, compared with the period before March 2020, was associated with a non-significant reduction in mean 25-hydroxyvitamin D (25(OH)D) concentration of 1.605 units (p=0.477). However, the interaction between the period after March 2020 and time was not significant (p=0.909), suggesting that there was no additional monthly change in 25-hydroxyvitamin D (25(OH)D) concentration after March 2020 beyond the increase. overall monthly. The constant term indicated a significant baseline mean 25-hydroxyvitamin D (25(OH)D) concentration of 24.004 units at the start of the study (p<0.001). The general model explained approximately 40.28% of the variation in the average monthly concentration of 25-hydroxyvitamin D (25(OH)D) (R-squared=0.4028, R-squared=0.3629) and was statistically significant (F=10.12, p <0.001).

**Table 2:** Regression analysis of monthly average 25-hydroxyvitamin D (25(OH)D) concentration, before and after March 2020.

| Variable | Coefficient |  |  |
| --- | --- | --- | --- |
|  | (Average Monthly 25-hydroxyvitamin D (25(OH)D) concentration) | t statistic | p-value |
| Time* (per each month) | 0.133 | 2.90 | 0.006 |
| Post-March 2020** (1) | -1.605 | -0.72 | 0.477 |
| Post-March 2020*Time*** | 0.008 | 0.11 | 0.909 |
| Constant**** | 24.004 | 33.94 | <0.001 |
| * The "Time" coefficient refers to the average monthly increase in 25-hydroxyvitamin D (25(OH)D) concentration across all participants in the study.<br>** "Post-March 2020 (1)" is the average difference in 25-hydroxyvitamin D (25(OH)D) concentration for all months after March 2020 (the 27th month of the study), compared to the months before March 2020.<br>*** "Post-March 2020*Time" is the interaction term, representing any additional monthly change in Vitamin D concentration after March 2020, beyond the general monthly increase.<br>****The Constant term represents the average 25-hydroxyvitamin D (25(OH)D) concentration at the start of the study (Time = 0). |  |  |  |

The proportion of toxicity across 2018 to 2021 increased significantly and the inadequate levels of 25-hydroxyvitamin D (25(OH)D) reduces in the same period, but with a non-significant change (**S3, S4 Tables)**. These findings highlight the presence of a general upward trend in 25-hydroxyvitamin D (25(OH)D) concentration over time, but no additional significant monthly change after the onset of the COVID-19 pandemic in March 2020. Results from the regression model Poisson tests indicate a significant increase in the prevalence of toxicity (**Table 3**) during the period from 2018 to 2021. Specifically, the prevalence of toxicity was found to be 6.53 times higher in 2020 and 7.37 times higher in 2021 compared to the base year 2018. In contrast, the prevalence of inadequate 25-hydroxyvitamin D (25(OH)D) levels (**Table 4**) showed a slight but not statistically significant decrease over the same period. The prevalence was found to be 3.5% lower in 2020 and 8% lower in 2021 compared to 2018. These findings suggest divergent trends in the two health outcomes over the study period.

**Table 3:**
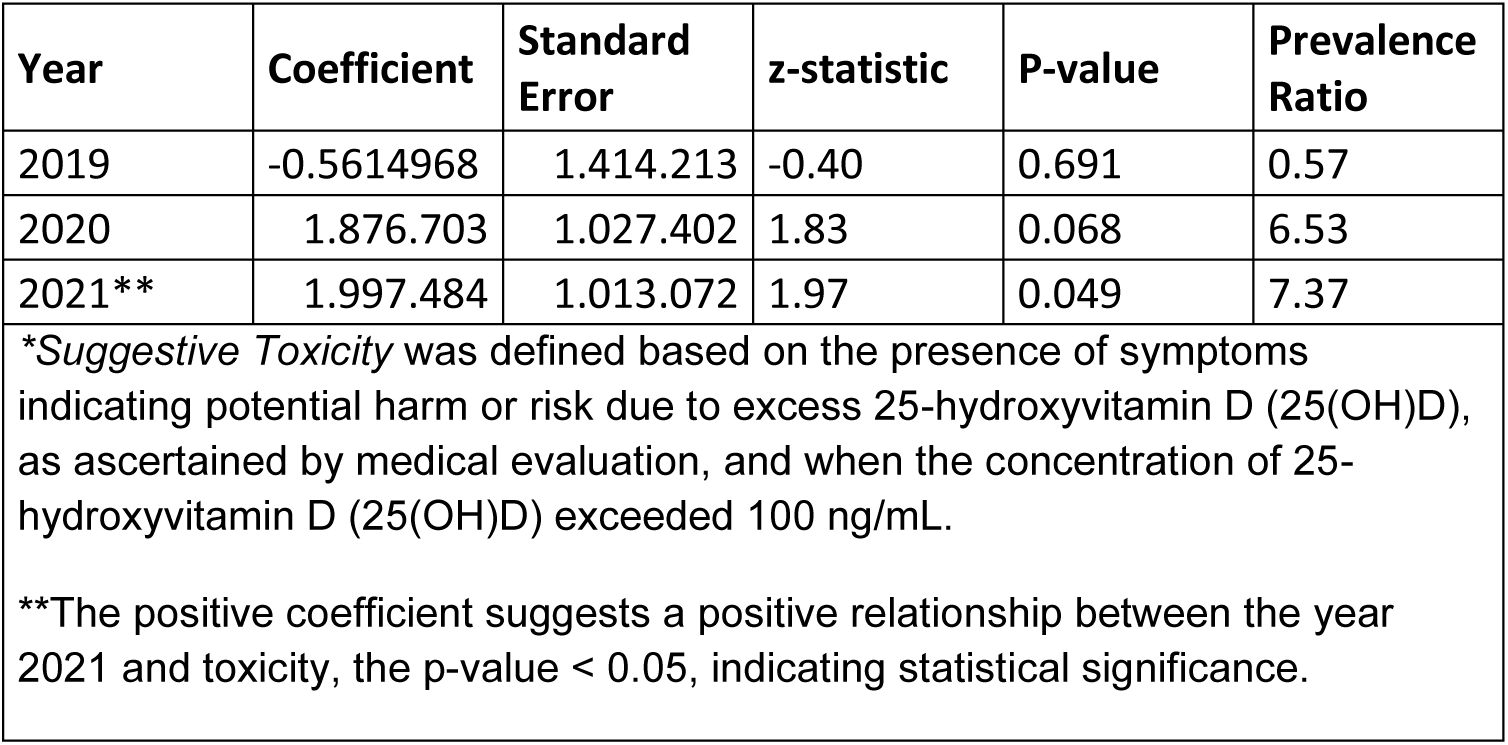
Poisson Regression Results for Toxicity* (2019-2021)

**Table 4:** Poisson Regression Results for Inadequate Vitamin D Level* (2019-2021)

| Year | Coefficient | Standard Error | z-statistic | P-value | Prevalence Ratio |
| --- | --- | --- | --- | --- | --- |
| 2019** | 0.0111135 | 0.0524749 | 0.21 | 0.832 | 1.01 |
| 2020** | -0.0359239 | 0.0492221 | -0.73 | 0.465 | 0.965 |
| 2021** | -0.0830686 | 0.0461695 | -1.80 | 0.072 | 0.92 |
| * *Inadequate 25-hydroxyvitamin D (25(OH)D) Level* was defined as having a concentration of 25-hydroxyvitamin D (25(OH)D) less than 20 ng/mL.<br>** These results suggest that there is no strong and consistent evidence for significant changes in the prevalence of inadequate vitamin D levels across the years 2019, 2020, and 2021. |  |  |  |  |  |

## Discussion

### Main findings

This study revealed a high prevalence of inadequate levels of 25-hydroxyvitamin D (25(OH)D) (64.58%) among individuals attending a private laboratory in Quito, Ecuador. These findings align with a meta-analysis conducted between 2000 and 2022, which estimated the global and regional prevalence of deficiency in serum 25-hydroxyvitamin D levels. The meta-analysis included 308 studies with 7,947,359 participants from 81 countries, finding that 15.7% had levels below 12 ng/dL and 47.9% had levels below 20 ng/dL. Although the prevalence slightly decreased from 2000–2010 to 2011–2022, it remained at a high level[10]. Similarly, another meta-analysis conducted in an Asian population, including 746,564 subjects, reported that 22.82% had levels below 12 ng/dL and 57.69% had levels below 20 ng/dL, indicating a high prevalence of 25-hydroxyvitamin D (25(OH)D) deficiency in Asia. 25-hydroxyvitamin D (25(OH)D) levels varied based on gender, age group, region, altitude, and disease. Several factors contributed to these results, including dietary habits. Vitamin D is only present in a few foods, such as fish liver oil and fatty fish, which are not commonly consumed by Asians, especially central Asians. Additionally, Asians tend to prefer a lighter skin color, leading to the use of sunscreen and parasols outdoors, which reduces the penetration of ultraviolet B (UVB) and subsequently decreases vitamin D synthesis. Clothing habits also play a significant role, as many individuals in western and southern Asia, where Muslim populations reside, wear long robes and veils that cover the skin. Moreover, people living in southern and Southeast Asia generally have darker skin, which can block UVB penetration. Economic development also influences vitamin D levels, particularly in Asian countries that are still developing and lack access to effective vitamin D supplements[10].

Furthermore, another meta-analysis estimated the prevalence of 25-hydroxyvitamin D (25(OH)D) deficiency in Africa. This analysis included 119 studies with 21,474 participants from 23 countries. The findings indicated that 18.46% had values below 12 ng/dL and 34.22% had values below 20 ng/dL. The prevalence of 25-hydroxyvitamin D (25(OH)D) deficiency exhibited regional variation, with the highest rates observed in northern African countries and South Africa[11]. Among population subgroups, women, newborn babies, and urban populations had the lowest concentrations of 25(OH)D. Notably, populations living in urban areas exhibited lower 25(OH)D concentrations compared to rural populations, potentially due to limited sunlight exposure duration or reduced dietary intake of vitamin D. Conversely, the populations in Africa that practiced traditional lifestyles, including nomadic animal rearing, hunting, and gathering, demonstrated the highest 25(OH)D concentrations[11].

In a meta-analysis that estimated the prevalence of 25-hydroxyvitamin D (25(OH)D) deficiency in South America, 96 studies with a total of 227,758 participants were included from an initial pool of 9,460 articles. The overall prevalence of 25-hydroxyvitamin D (25(OH)D) deficiency, defined by a 25(OH)D concentration below 20 ng/mL, was found to be 34.76%. Lifestyle factors such as spending more time indoors for work, leisure, and physical activities, coupled with dietary patterns and public health campaigns that promote sunlight avoidance and skin protection, likely contribute to the lower-than-expected concentrations of 25(OH)D in this region[5].

Ecuador, situated in the tropical zone, is longitudinally crossed by the Andes mountain range, which imparts distinct and prominent topographical characteristics throughout the country. The altitude of Ecuador’s regions holds significant influence over various health outcomes experienced by its population[12]. Within the scope of this study, a pronounced disparity in inadequate 25-hydroxyvitamin D (25(OH)D) levels was observed in laboratory results obtained from individuals residing in Quito, positioned at an approximate elevation of 2,850 meters above sea level, in comparison to samples collected from individuals in Santo Domingo, situated at a lower altitude of approximately 625 meters above sea level. Upon comparing participants from Quito, Ambato, and Ibarra with those from Santo Domingo, it was found that 64.18% of individuals from higher-altitude cities exhibited inadequate 25-hydroxyvitamin D (25(OH)D) levels, whereas the percentage was 52.74% among those from Santo Domingo. These findings indicate a potential correlation between altitude and 25-hydroxyvitamin D (25(OH)D) status, suggesting that individuals inhabiting higher altitudes are more susceptible to insufficient levels of this nutrient. Therefore, it is crucial to consider regional factors when evaluating 25-hydroxyvitamin D (25(OH)D) status and devising appropriate interventions. These results align with other studies, including research conducted in Asia by Zhiwei Jiang, which revealed that individuals residing at lower altitudes (≤500 m) tend to exhibit higher 25-hydroxyvitamin D (25(OH)D) levels compared to those at higher altitudes (>500 m)[10]. Furthermore, it has been observed that people living in high-altitude regions often encounter difficulties in accessing effective vitamin D supplements[10]. The studies conducted in South America have incorporated two fundamental factors, namely altitude and diet, into their analyses. At higher altitudes, the atmosphere is thinner, resulting in reduced absorption of ultraviolet radiation (UV levels increase by 10% to 12% with each 1000-meter increase in altitude)[5].

The findings of this study revealed a significant prevalence of inadequate 25-hydroxyvitamin D (25(OH)D) levels in the population, while the incidence of 25-hydroxyvitamin D (25(OH)D) toxicity was relatively low (0.62%). Interestingly, divergent changes were observed in these indicators during the study period. Although inadequate levels showed a non-significant reduction, there was a notable increase in the prevalence of toxicity from 2018 to 2021. These results underscore a concerning trend in the 25-hydroxyvitamin D (25(OH)D) status of the population. These findings are consistent with a study conducted in Ireland that aimed to investigate the impact of the COVID-19 pandemic on vitamin D status and the usage of newly introduced vitamin D supplements. A trend analysis based on laboratory data revealed a threefold rise in the yearly average of 25(OH)D during the initial year of the pandemic, compared to previous analyses. This trend suggests potential benefits for individuals with low vitamin D status but also highlights the risk for those with already high levels, especially considering the increasing availability of high-dose supplements[6].

The increased consumption of vitamin D supplements by the general population, including therapeutic and high-dose formulations, without adequate medical supervision, can significantly elevate the risk of exogenous hypervitaminosis D, commonly known as vitamin D toxicity. This condition can manifest symptoms of hypercalcemia[13]. Therefore, it is essential to ensure that the intake of vitamin D is monitored by healthcare professionals to prevent overdosing and the associated severe health consequences[3,14,15].

Excessive intake of vitamin D can lead to the accumulation of the nutrient in the body for extended periods, up to 18 months, resulting in chronic toxic effects such as nephrocalcinosis, hypercalcemia, and hypercalciuria. In the past, fortification of foods, such as milk, was recommended as a public health strategy to prevent vitamin D deficiency and low 25-hydroxyvitamin D (25(OH)D) status. However, cases of increased hypercalcemia associated with excessive intake of fortified foods have been reported[3,14,15].

During the pandemic, there was a notable interest in studying the relationship between 25-hydroxyvitamin D (25(OH)D) levels and COVID-19. A study aimed at identifying if online search interest in vitamin D increased with the pandemic burden and analyzing the accuracy of public health messaging regarding vitamin D in online news articles found that a significant number of articles provided conflicting information or incorrectly advised supratherapeutic doses. This study emphasizes the opportunity for public health organizations to capitalize on the increased interest in vitamin D during the pandemic and disseminate accurate information to raise awareness[16].

However, it is important to acknowledge the limitations of this study. Important factors such as skin pigmentation, socioeconomic conditions, diet, sun exposure habits, cultural practices, and skin coverage with clothing were not considered in the analysis. These factors can influence 25-hydroxyvitamin D (25(OH)D) levels and should be considered in future research.

The results of this study reveal a high prevalence of 25-hydroxyvitamin D (25(OH)D) deficiency, indicating the need for strategies to prevent deficiency. However, the increase in toxicity also raises concerns and highlights the importance of rational prescribing of vitamin D supplements and educating the population to avoid self-medication. To optimize resource allocation and prioritize those at higher risk, it may be beneficial to focus vitamin D testing on specific populations. Individuals with malabsorption syndromes, individuals undergoing steroid therapy, or older adults who are confined to their homes are examples of higher-risk groups. Assessing serum of 25-hydroxyvitamin D (25(OH)D) levels in these cases can provide valuable clinical insights and inform appropriate interventions.

## Data Availability

All data produced in the present study are available upon reasonable request to the authors.

## Acknowledgment

The authors express their sincere appreciation to the healthcare professionals at Zurita & Zurita Laboratories for their expert handling of the samples, which significantly contributed to the accuracy of our study. Furthermore, ChatGPT from OpenAI was utilized to improve the document’s overall writing quality. No AI was involved in the study design, data analysis, interpretation or substantial writing and reviewing of the manuscript.

## Funding statement

This research received no specific grant from any funding agency in the public, commercial or not-for-profit sectors.

## Author Contributions

Conceptualization: Camilo Zurita-Salinas, Betzabé Tello, Iván Dueñas-Espín

Data curation: Iván Dueñas-Espín, Betzabé Tello, Camilo Zurita-Salinas

Formal analysis: Iván Dueñas-Espín, Betzabé Tello, Camilo Zurita-Salinas

Investigation: Camilo Zurita-Salinas, Betzabé Tello, Iván Dueñas-Espín, Jeannete Zurita, William Acosta, Cristina Aguilera León, Andrés Andrade-Muñoz, José Pareja-Maldonado.

Methodology: Camilo Zurita-Salinas, Betzabé Tello, Iván Dueñas-Espín

Validation: Camilo Zurita-Salinas, Betzabé Tello, Iván Dueñas-Espín, Jeannete Zurita, William Acosta, Cristina Aguilera León, Andrés Andrade-Muñoz, José Pareja-Maldonado.

Writing – original draft: Betzabé Tello, Iván Dueñas-Espín

Writing – review & editing: Camilo Zurita-Salinas, Betzabé Tello, Iván Dueñas-Espín, Jeannete Zurita, William Acosta, Cristina Aguilera León, Andrés Andrade-Muñoz, José Pareja-Maldonado.

## Declaration of interest

None

## Patient consent for publication

Not required.

## Data-sharing statement

Data are available upon reasonable request. All data relevant to the study were included in the article.

---

**S1 Table:**
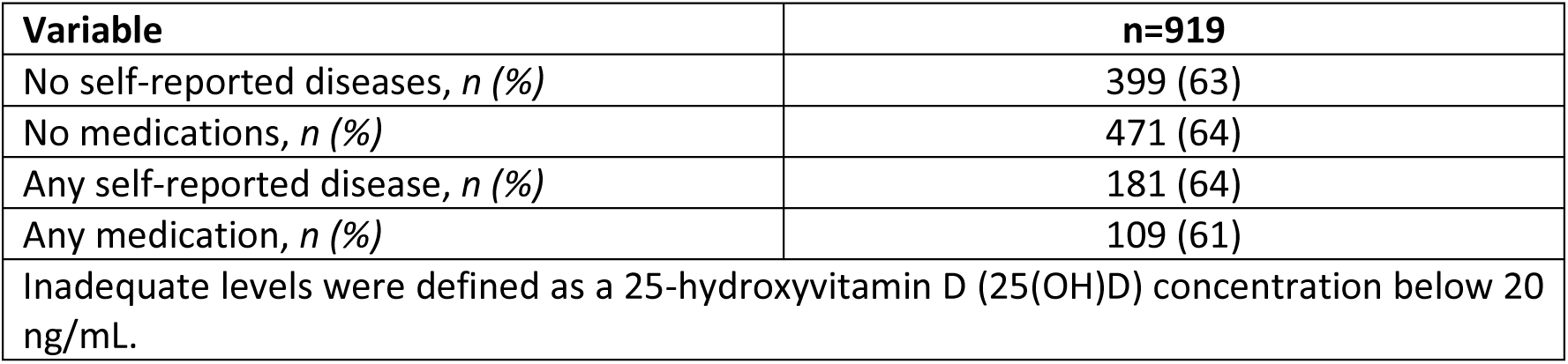
Prevalence of inadequate levels of Vitamin D among: (i) those who does not have self-reported diseases, (ii) those who are not taking medications, (iii) those who have reported any chronic disease, and (iv) those who are taking any medication.

**S2 Table:**
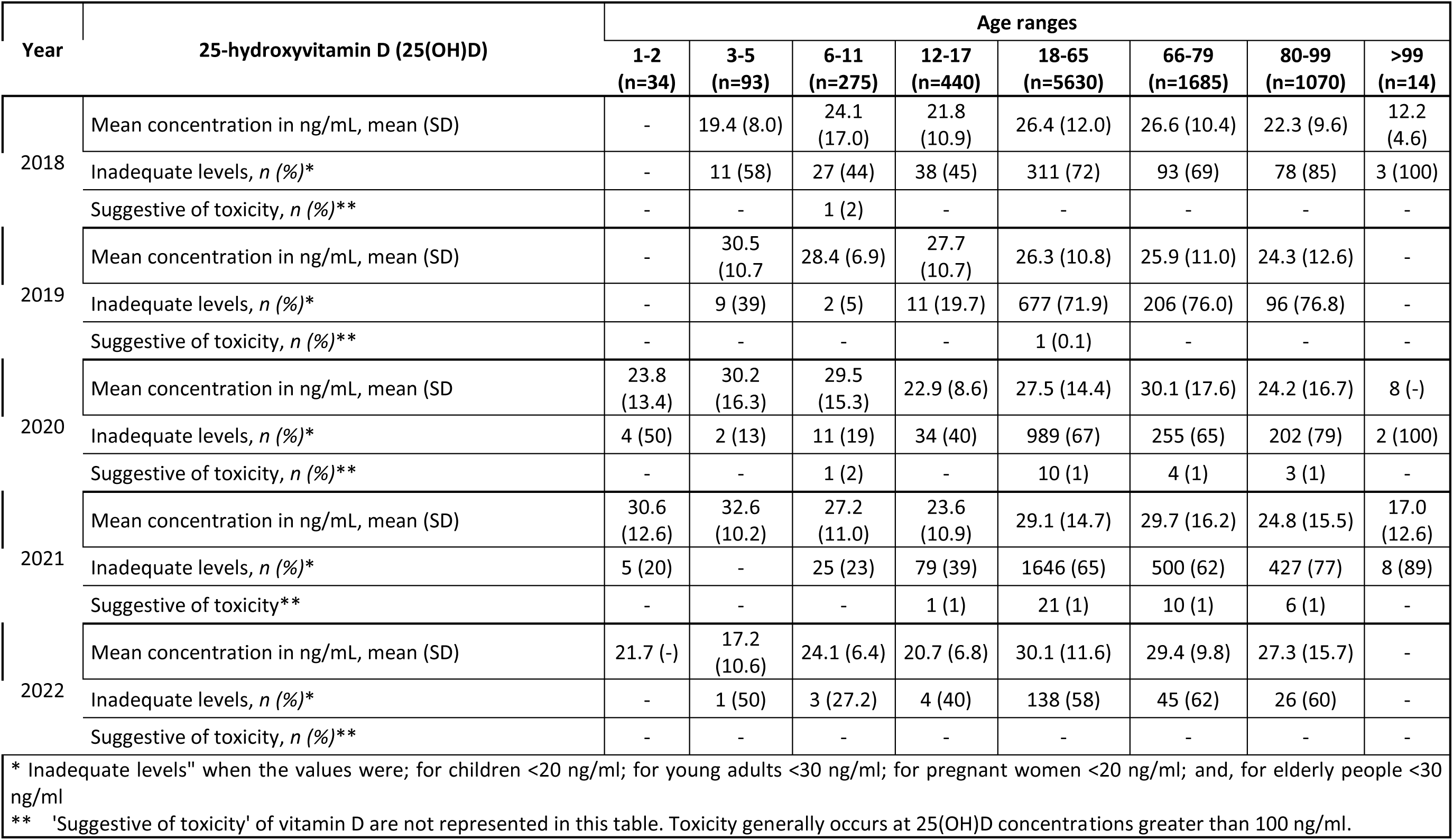
25-hydroxyvitamin D (25(OH)D) levels, suggestive toxicity, and inadequate levels per year and age group.

**S3 Table:** Annual Prevalence Rates of Vitamin D Toxicity (%).

| Year | n | Prevalence Rate (%) | 95% Confidence Interval |
| --- | --- | --- | --- |
| 2018 | 835 | 0.12 | 0.02 - 0.85 |
| 2019 | 1464 | 0.07 | 0.01 - 0.48 |
| 2020 | 2301 | 0.78 | 0.49 - 1.24 |
| 2021 | 4305 | 0.88 | 0.64 - 1.21 |
| Toxicity generally occurs at 25(OH)D concentrations greater than 100 ng/ml. |  |  |  |

**S4 Table:** Proportion of patients with inadequate vitamin D levels per year.

| Año | n | Proporción | Intervalo de confianza del 95% |
| --- | --- | --- | --- |
| 2018 | 835 | 0.68 | 0.65 - 0.71 |
| 2019 | 1464 | 0.69 | 0.66 - 0.71 |
| 2020 | 2301 | 0.66 | 0.64 - 0.68 |
| 2021 | 4305 | 0.63 | 0.61 - 0.64 |
| Less than 20 ng/ml for general population and less than 30 ng/ml for older adults and pregnant women |  |  |  |

## Notes

### Competing Interest Statement

The authors have declared no competing interest.

### Funding Statement

This study did not receive any funding.

### Summary of Updates

Updated authorship order for the article

